# Role of Heterogeneous Transmission in the Decline of COVID-19 Cases During Winter of 2020/2021 in Massachusetts

**DOI:** 10.1101/2021.02.18.21251955

**Authors:** Yeon-Woo Choi, Marcia C. Castro, Elfatih A. B. Eltahir

## Abstract

**Importance:** Heterogeneity in transmission of COVID-19 is a significant multiscale phenomenon. However, the role of this heterogeneity in shaping the overall dynamics of disease transmission is not well understood.

**Objective:** To investigate the role of heterogeneous transmission among different towns in Massachusetts in shaping the dynamics of COVID-19 transmission, especially the recent decline during winter of 2020/2021.

**Design, Setting, Participants:** Analysis of COVID-19 data collected and archived by the Massachusetts Department of Public Health.

**Exposures:** The entire population of the state of Massachusetts is exposed to the virus responsible for COVID-19, to varying degrees. This study quantifies this variation.

**Main outcome measures:** Weekly observations, by town, on confirmed COVID-19 cases in Massachusetts, during the period (April 15^th^, 2020 to February 9^th^ 2021).

**Results:** The relative decline in COVID-19 cases, during January 12^th^, 2021 to February 9^th^, 2021, in the group of towns with higher total accumulated cases in the period before January 12^th^, 2021 is significantly larger than the corresponding relative decline in the group of towns with lower accumulated cases during the same period.

**Conclusions and Relevance:** Heterogeneous nature of transmission is playing a significant role in shaping the rapid recent decline (January 12^th^ to February 9^th^, 2021) in reported cases in Massachusetts, and probably around the country. These findings are relevant to how we estimate the threshold defining “herd” immunity, suggesting that we should account for effects due to heterogeneity.

**Key Points:** *Question:* Does heterogeneity in disease transmission play a role in shaping the overall dynamics of COVID-19 in Massachusetts, including the recent decline in cases during the 2020/2021 winter.

*Findings:* Based on analysis of data on cases in Massachusetts, the consistent and widespread decline of COVID-19 spread during winter of 2020/2021 (January 12^th^, 2021 to February 9^th^, 2021) appears to be shaped to a significant degree by the heterogeneous nature of transmission at the scale of different towns. Towns with a history of high (low) transmission rates during 2020 are experiencing a faster (slower) relative decline.

*Meaning:* We suggest that heterogeneity in transmission of COVID-19 may impact the dynamics of disease transmission including the emergence of “herd” immunity, in line with some recent theoretical studies. This finding deserves some attention from other research groups investigating “herd” immunity, and from federal and state public health authorities concerned with the future evolution of the pandemic.

## Introduction

The intensity of transmission of the novel coronavirus disease 2019 (COVID-19) varies significantly across towns of Massachusetts (MA). Figure 1 shows the frequency distribution of the total accumulated COVID-19 cases before the January 12^th^, 2021 peak (hereafter referred to as accumulated cases), and their spatial distribution. The mean of accumulated cases is 44.5 per thousand people and the standard deviation is 26 per thousand indicating significant heterogeneity in the transmission of the disease.

**Figure 1.**
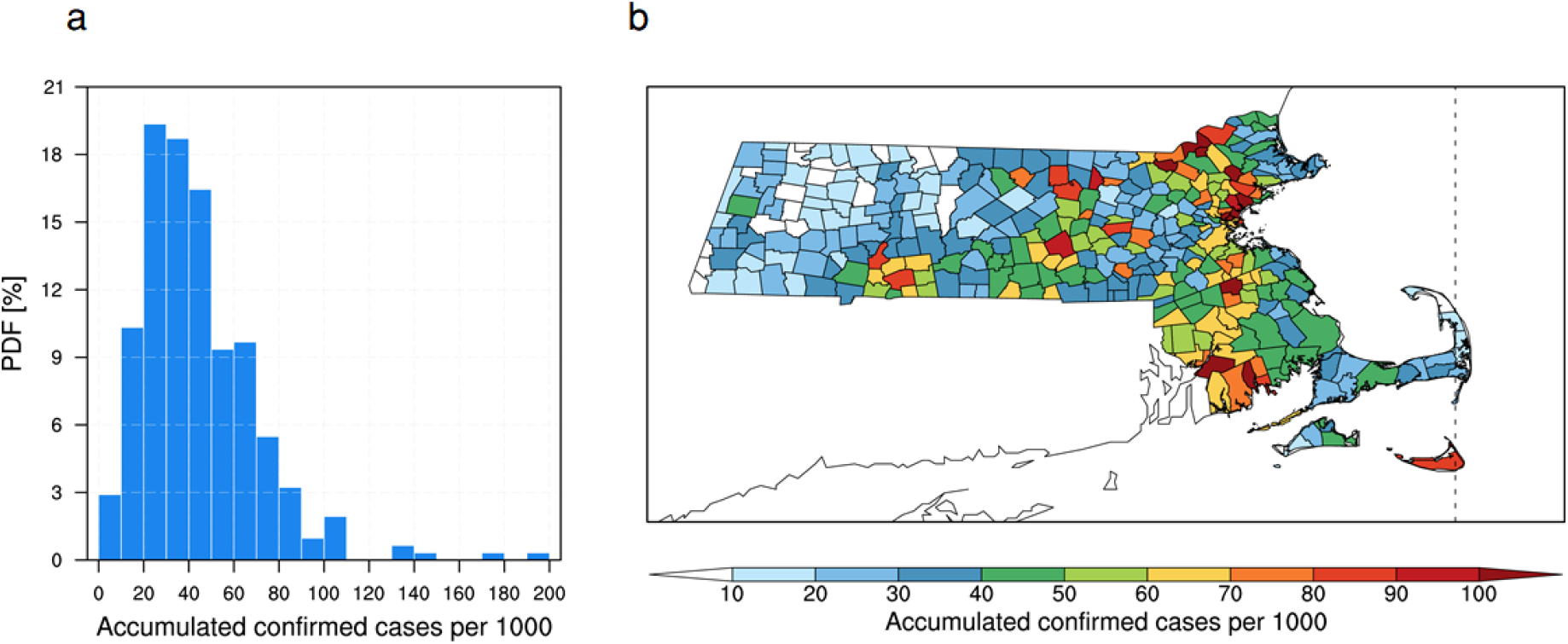
Accumulated confirmed COVID-19 cases, before January 12^th^ 2021, in Massachusetts. Probability density function (a) and spatial distribution (b) of accumulated confirmed COVID-19 cases per 1,000 people before January 12th, 2021, across towns within the state of Massachusetts.

The SARS-CoV-2 virus responsible for COVID-19 was initially reported in Wuhan, China,^1,2,3^ and it rapidly spread to various regions of the Northern Hemisphere at the early epidemic stage.^4^ The first infection by SARS-CoV-2 in MA was detected by the state public health department in early February 2020. Since then, a medical conference that took place on February 26-27 in Cambridge, MA, caused the outbreak to surge.^5^ Despite government intervention measures, the virus has been rapidly spreading throughout the state beyond Cambridge, peaking at the end of April.^6^ The state faced a second wave of the COVID-19 pandemic that started in November 2020, causing a total of 521,045 confirmed cases, including 14,903 associated deaths as of February 9^th^, 2021. The number of confirmed cases has been declining after a second peak around January 12^th^, 2021.^7^

Recent modeling and theoretical studies proposed that heterogeneity in transmission can decrease the herd immunity threshold.^8,9^ In particular, variation in individual susceptibility to infection may significantly decrease the threshold of herd immunity (to less than 60%). According to classical herd immunity theory applied to COVID-19 pandemic, it is estimated that 60-80% of a homogeneous population have to become immune to reach herd immunity, assuming a reproductive number (R0) between 2.5 and 5.^10,11,12^ However, this classical approach cannot apply to infection-induced immunity in real-world situations since natural infections do not occur randomly, and R0 is not uniform across the population.^9,13^ Although it has been suggested that the herd immunity threshold could be lowered due to heterogeneity, there is no agreement on the magnitude of the threshold because scarce epidemiological data for this pandemic has not been able to provide definitive observation-based evidence.

The recent consistent and widespread decline of COVID-19 prevalence in MA, as well as the USA, gives us a strong motivation and context for the investigation of heterogeneous disease dynamics. The main objective of this study is to investigate the role of heterogeneous transmission in shaping the dynamics of COVID-19 in MA, especially the significant decline during winter of 2020/2021.

## Methods

### Data Collection

Weekly data on confirmed COVID-19 cases, number of tests, and positivity rate (i.e., total number of positive test as a fraction of the total number of tests) at the scale of different towns within the state of MA, for the period April 15^th^, 2020 to February 9^th^, 2021, are accessed from an archive of COVID-19 weekly public health reports (available at https://www.mass.gov/info-details/archive-of-covid-19-weekly-public-health-reports). The data for April 15^th^ 2020 includes accumulated cases before that date. We applied a count of confirmed cases per one thousand people to assess the overall impact of a COVID-19 pandemic on towns of different population sizes in MA. Among a total of 351 towns in MA, this study considered 310 towns where total accumulated confirmed COVID-19 cases on January 12^th^ 2021 are not zero and there are no missing values during the analysis period.

Town-level population data are taken from the annual estimates of the resident population for incorporated places in Massachusetts.^14^ With this data, the density of population is calculated by dividing the population by area which is available at https://en.wikipedia.org/wiki/List_of_municipalities_in_Massachusetts. To identify the potential effect of poverty on COVID-19 spread, median house income data is utilized, which is available at https://en.wikipedia.org/wiki/List_of_Massachusetts_locations_by_per_capita_income. The results for this analysis are shown in Supplementary Material.

### Statistical Analysis

We investigate heterogeneity in transmission of COVID-19 at the scale of different towns within the state of Massachusetts. The analyses are conducted for the period from April 15^th^, 2020 to February 9^th^, 2021 by considering and comparing different groups: (1) all towns in MA (i.e., 310 towns are considered; see Data Collection); (2) two groups, each with the same number of towns; (3) two groups, each with the same number of people of about 3.3 million; and (4) two groups each with the same number of people of about 1 million. Time series analysis is presented to identify recent trend of COVID-19 incidence. Decreasing pattern of weekly COVID-19 cases as a function of total accumulated cases is presented, across the towns, for a period of about one month (January 12^th^, 2021 to February 9^th^, 2021).

We perform “bootstrap” analysis, resampling without replacement, (repeated 1,000 times) to check statistical significance of the difference in relative decline during the period (January 12^th^, 2021 to February 9^th^, 2021) between the two groups of towns with high and low accumulated COVID-19 confirmed cases.

Statistical relationships between accumulated confirmed COVID-19 cases on January 12^th^, 2021 and several independent predictors (i.e., population density, and median house income) are developed using correlation analyses and presented in scatter plots (See Supplementary Material). *p* values are calculated using two-sided tests.

### Analysis of Heterogeneity

We use the example of MA, whose recent trend is shown in Figure 2a (January 12^th^ to February 9^th^, 2021), to analyze the variability in the dynamics of the disease across towns. We seek to determine how and why the magnitude of the decline varied across the different towns. In a homogeneous transmission mode, the magnitude of the decline would be uniform across the different towns. However, given the heterogeneous nature of the transmission in this state, and across the country, we would like to understand the origin and implications of this heterogeneity, and in particular if the associated variation in previous infection burden plays any role. This last point is important since we expect the distribution of immunity to reflect the heterogeneous transmission pattern. We will assume that exposure to the virus as well as level of acquired immunity are directly proportional to the accumulated cases. The more cases reported, the higher the level of immunity among the population.

**Figure 2.**
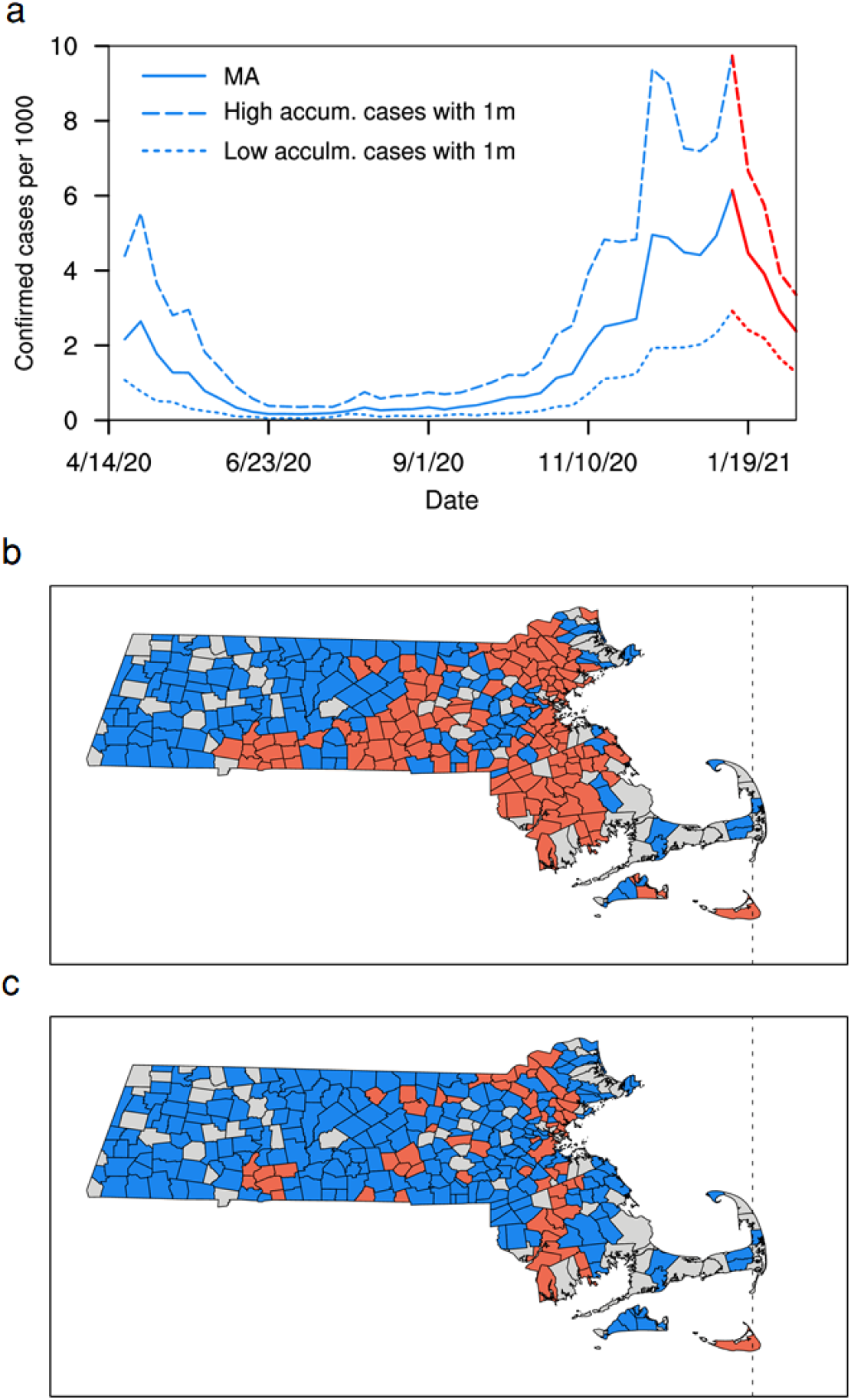
Temporal and spatial patterns of COVID-19 outbreak (a). Times series of confirmed COVID-19 cases averaged over Massachusetts (thick line), and averaged over two groups of towns, each group with 1 million people representing towns with high (dashed line) and low (dotted line) accumulated confirmed cases. Spatial distribution of two groups each with the same number of towns (b). Spatial distribution of two groups each with the same number of people of about 3.3 million (c). Red and blue colours indicate towns with high and low cumulative confirmed COVID-19 cases, respectively. Grey colour indicates towns with missing data points.

## Results

### Implications of Heterogeneity to Disease Transmission Dynamics

Starting around the 12^th^ of January 2021, the daily cases of COVID19 in MA started to decline significantly (Figure 2a). In about one month, the daily cases declined by 61% from 6.1 cases per thousand to 2.4 per thousand. This large decline reflects a similar trend in cases documented in the USA, with declines reported in most of the States, and an overall rate of decline larger than that of any downward trends since the emergence of this disease.

We estimate that 92% of all the towns reported a decline in cases, 8% of the towns reported an increase in cases during the same period, Table 1. When Massachusetts towns are sorted according to accumulated cases, and then split into two groups each with the same number of towns, a pattern emerges. For towns with low accumulation, 15% of the towns reported an increase, while 85% reported a decline. For towns with relatively high accumulation, 1% of the towns reported an increase, while 99% reported a decline. This finding indicates that towns with relatively high accumulated cases are more (less) likely to show a decline (an increase) in cases compared to towns with relatively low accumulated cases. However, the size of the population in the two groups of towns is not equal, 5.1 million for low accumulated cases versus 1.5 million for high accumulated cases.

**Table 1.** Towns with increasing and decreasing trends of confirmed COVID-19 cases (January 12^th^ 2021 to February 9^th^, 2021). Two groups each with the same number of towns (top three lines). Two groups each with the same number of people of about 3.3 million (bottom three lines). Total of 310 towns are considered where accumulated confirmed COVID-19 cases on January 12, 2021 are not zero and there are no missing values during the analysis period.

|  | **Towns with low accumulated cases (155 towns, 1,506,050 people)** | **Towns with high accumulated cases (155 towns, 5,073,330 people)** |
| --- | --- | --- |
| **Number (percent) of town with increasing trend** | 24 (15%) | 2 (1%) |
| **Number (percent) of town with decreasing trend** | 131 (85%) | 153 (99%) |
|  | **Towns with low accumulated cases (241 towns, 3,297,540 people)** | **Towns with high accumulated cases (69 towns, 3,281,840 people)** |
| **Number (percent) of town with increasing trend** | 25 (10%) | 1 (1%) |
| **Number (percent) of town with decreasing trend** | 216 (90%) | 68 (99%) |

When towns are sorted according to accumulated cases and split into two groups each with the same number of people of about 3.3 million, the pattern identified above persists. Defining the groups this way shifts the accumulated cases threshold from 40 cases per thousand to 61 cases per thousand. For towns with low accumulation, 10% of the towns reported an increase, while 90% reported a decline. For towns with relatively high accumulation, 1% of the towns reported an increase, while 99% reported a decline. This finding confirms that towns with relatively high accumulated cases are more (less) likely to show a decline (an increase) in cases compared to towns with relatively low accumulated cases. The patterns shown in Figure 2 (b and c) identifies these towns, at different thresholds of accumulated cases, 40 versus 61 cases per thousand.

The number of cases reported in any town for the week of January 12^th^ 2021 is directly proportional to the total number of accumulated cases for that town. This relationship is shown in Figure 3 (a), indicating a significant correlation between the two variables of about 0.8. If the dynamics of disease transmission, as characterized by a variable such as the reproduction number, is of similar magnitudes among the towns, you would expect theoretically a change in cases during the recent period proportional to the reported cases on January 12^th^, 2021, which is directly proportional to the accumulated cases in each town, according to Figure 3a. Figure 3 (b and c) show significant declines in observed cases and in positivity that are proportional to the accumulated cases. Towns with larger accumulated cases are reporting larger declines during the month following January 12^th^, 2021. Consistent with the observed relationships presented in Figure 3, the average confirmed cases for the week of January 12^th^ 2021 in the group of towns with high accumulated cases is about 6.9 cases per thousand, compared to 3.5 cases per thousand for the lower accumulated cases group, with the same population size (Figure 4a). Similarly, the declines for the two groups of towns are roughly 4.3 and 2 cases per thousand, over the period (January 12^th^ to February 9^th^, 2021). If the reproduction number in the two groups of towns are identical you would expect, that the relative change in the cases among the two groups of towns would be the same. However, Figure 4b shows a relative decline in cases in the group of towns with higher accumulated cases that is significantly larger than the corresponding relative decline in the group of towns with lower accumulated cases. If we use the relative magnitude of cases at the end of the period (defined as the fraction of cases at the end of the period normalized by the number of cases at the beginning of the period) as an integrated proxy for the reproduction number, then the comparison in Figure 4b indicates that our proxy for reproduction number is smaller in the group of towns with higher accumulated cases in comparison to the same variable in the group of towns with lower accumulated cases. These results are based on the data for two groups of towns, each group with the same population of about 3.3 million. Similar analysis is carried for two groups of towns, each with a population of 1 million, representing the two ends of the distribution of accumulated cases (Figure 4c and d). The corresponding time series of cases for the two groups of towns were shown in Figure 2a. The results are similar but the contrast is sharper, indicating a difference in our proxy reproduction number by a factor of about 1.3 (significant at 1% level), from a fraction of 34% by the end of the period to a fraction of 44% by the end of the period, (January 12^th^ to February 9^th^, 2021). This significant difference between the two groups of towns indicates a significant impact of the history of accumulated cases in the COVID-19 transmission dynamics.

**Figure 3.**
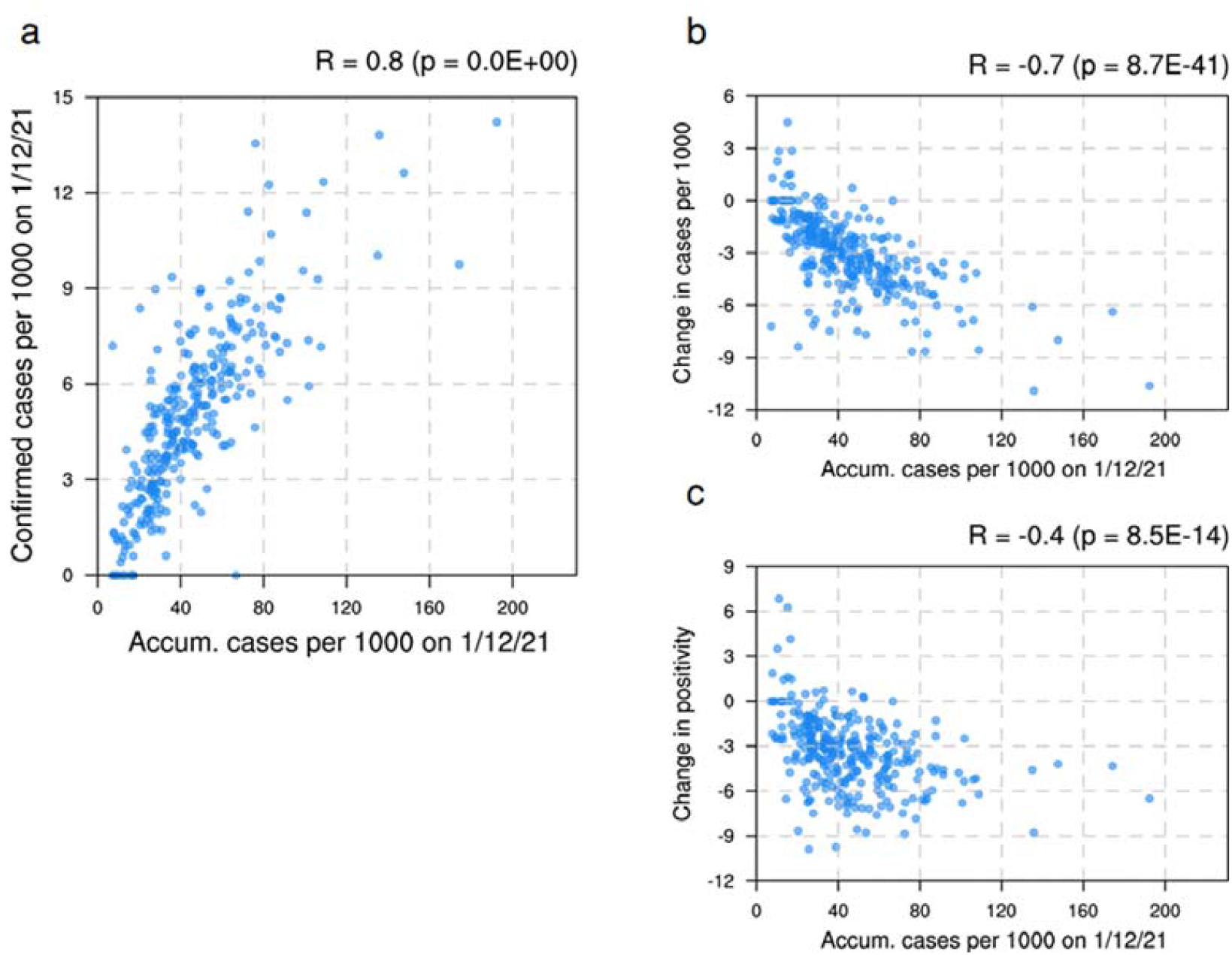
Confirmed COVID-19 cases across towns within the state of Massachusetts. Accumulated confirmed COVID-19 cases per 1,000 people before January 12^th^, 2021, against weekly confirmed cases per 1,000 people for January 12^th^ 2021 (a); change in confirmed cases per 1,000 people (b), and change in positivity in percent (c). Changes in b and c are estimated for the period from January 12^th^ to February 9^th^, 2021. Values indicated on top of each plot represent the correlation coefficient (R) and p-value (p). For (c), we exclude 8 towns where the recent number of tests are too low relative to the tests in the rest of the 310 towns.

**Figure 4.**
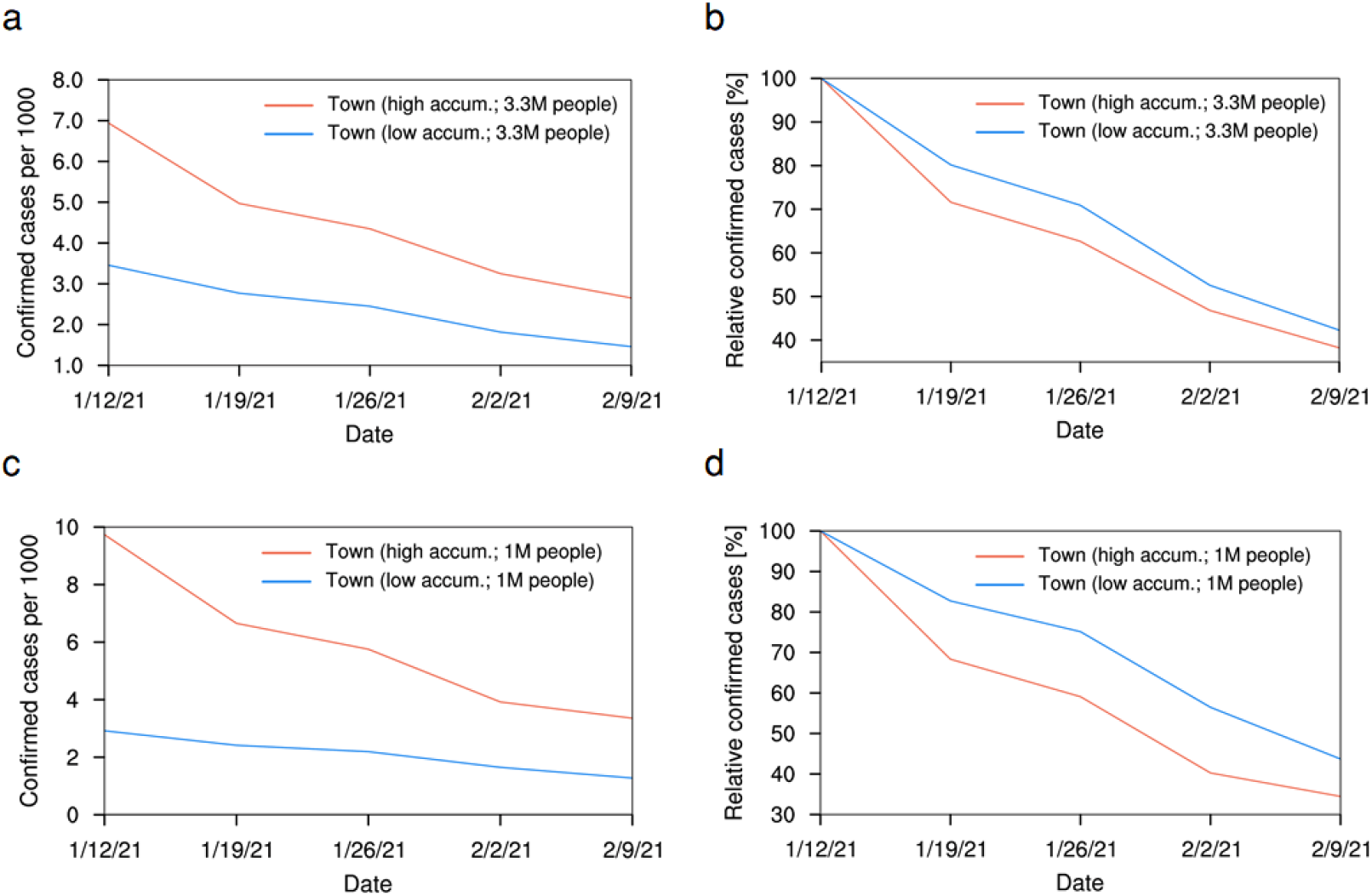
Trend of confirmed COVID-19 cases. Time series of weekly confirmed COVID-19 cases per 1,000 (a) and relative confirmed cases in percent (relative to cases on January 12^th^ 2021) (b) averaged over towns of 3.3 million people with high (red) and low (blue) accumulated confirmed cases. (c-d) same as (a-b), but for two groups of towns each with 1 million people, representing both ends of the distribution of Figure 1a.

## Discussion and Conclusions

Heterogeneity in disease transmission is likely to manifest itself at different scales. From the scale of a single-family, different members may have different potential exposures to the virus due to their different age-related habits, modes of mobility, and patterns of social mixing. Different neighborhoods within the same town may experience different intensities of transmission depending on housing types and population density. Different states and regions within the same country may experience different climates, and a different mix of urban/rural population, and as a result have different potential for disease transmission. Hence, in general, heterogeneity in transmission of COVID-19 is a multiscale phenomenon. Here, we only considered heterogeneity in disease transmission at the scale of different towns within the state of MA.

Our results indicate that heterogeneity at the scale of different towns is likely to play a significant role in shaping the dynamics of COVID-19 transmission. Our proxy for the reproduction number is smaller than one, and significantly smaller for the group of towns with higher accumulated cases compared to the group of towns with lower accumulated cases. Mechanistically, this finding can be explained as due to differences in social behavior between the two groups of towns. However, we are not aware of any documented differences. A more likely explanation is the difference in accumulated cases between the two different groups of towns. Towns with higher accumulated cases are locations where the virus circulation among the population has been relatively higher during the last year. We expect that the population experiencing such conditions would develop higher levels of immunity against further infections by the virus. If this is true, then the reduction in our proxy of reproduction number in the towns with higher accumulated cases would reflect the higher immunity levels in the population of those towns in comparison to the immunity levels in the population of towns with a record of lower accumulated cases. Unfortunately, there are no available direct observations on immunity levels among the population that may support such a hypothesis.

Our findings suggest that the heterogeneous nature of COVID-19 transmission is playing a significant role in shaping the rapid recent decline in reported cases in MA, and probably around the country. One of the scenarios for the elimination of COVID-19 from any population assumes that the level of natural immunity would increase to reach a threshold where new infections would be hampered and would occur at a rate lower than the rate of patients’ recovery. Under such conditions, new transmission is increasingly made impossible, leading to elimination of the virus. This threshold defines “herd” immunity. Estimates for herd immunity in the US population, as communicated by federal public health officials, range from 60 to 80%. There have been suggestions, however, that due to heterogeneous nature of the transmission herd immunity may be achieved at significantly smaller levels of immunity of about 40%.^8^ Most of these proposals were based on theoretical studies. Our findings, based on analysis of data on reported cases, support the idea that heterogeneous transmission can be an important factor in shaping the dynamics of COVID-19 transmission. Therefore, we recommend that lower estimates of the threshold for “herd” immunity, suggested by some recent theoretical studies, may deserve further investigation by other research groups investigating “herd” immunity, and some attention from federal and state public health authorities concerned with the future evolution of the pandemic.

## Data Availability

Weekly data on confirmed COVID-19 cases, number of tests, and positivity rate (i.e., total number of positive test as a fraction of the total number of tests) at the scale of different towns within the state of MA, for the period April 15th, 2020 to February 9th, 2021, are accessed from an archive of COVID-19 weekly public health reports (available at https://www.mass.gov/info-details/archive-of-covid-19-weekly-public-health-reports). The data for April 15th 2020 includes accumulated cases before that date. We applied a count of confirmed cases per one thousand people to assess the overall impact of a COVID-19 pandemic on towns of different population sizes in MA. Among a total of 351 towns in MA, this study considered 310 towns where total accumulated confirmed COVID-19 cases on January 12th 2021 are not zero and there are no missing values during the analysis period.
Town-level population data are taken from the annual estimates of the resident population for incorporated places in Massachusetts.14 With this data, the density of population is calculated by dividing the population by area which is available at https://en.wikipedia.org/wiki/List_of_municipalities_in_Massachusetts. To identify the potential effect of poverty on COVID-19 spread, median house income data is utilized, which is available at https://en.wikipedia.org/wiki/List_of_Massachusetts_locations_by_per_capita_income. The results for this analysis are shown in Supplementary Material.

https://www.mass.gov/info-details/archive-of-covid-19-weekly-public-health-reports

https://en.wikipedia.org/wiki/List_of_Massachusetts_locations_by_per_capita_income

https://en.wikipedia.org/wiki/List_of_Massachusetts_locations_by_per_capita_income

## Author Contributions

E. A. B. E. conceived and supervised the study. E. A. B. E., and M. C. C. contributed to the conceptual design of the study. Y. C. carried out analyses. E. A. B. E., M. C. C., and Y. C. contributed to the drafting and editing of the manuscript.

## Conflict of Interest Disclosures

None reported.

## Funding/Support

Breene M Kerr Chair at MIT.

## Role of the Funder/Sponsor

None reported.

## Disclaimer

The findings and conclusions in this report are those of the authors.

## Notes

### Competing Interest Statement

The authors have declared no competing interest.

