## Supplementary Material for "Role of Heterogeneous Transmission in the Decline of COVID-19 Cases During Winter of 2020/2021 in Massachusetts"

### Supplementary Information for
### "Role of Heterogeneous Transmission in the Decline of COVID-19 Cases During Winter of 2020/2021 in Massachusetts"

February 17, 2021

Authors:

Yeon-Woo Choi[1], PhD

Marcia C. Castro[2], PhD

Elfatih A. B. Eltahir[1,*], ScD

Affiliations:

[1]Ralph M. Parsons Laboratory, Massachusetts Institute of Technology, Cambridge, Massachusetts 02139, USA

[2]Department of Global Health and Population, Harvard T.H. Chan School of Public Health, Boston, Massachusetts, 02115, USA

Ralph M. Parsons Laboratory, Massachusetts Institute of Technology, Cambridge, Massachusetts 02139, USA

**Drivers of Heterogeneity in COVID-19 Transmission in Massachusetts**

**Introduction**

Recently, much attention is focused on the association of socio-demographic factors (e.g., age, gender, ethnicity, education level, economic, and occupational factors) with COVID-19 spread in Massachusetts at the sub-state scale. Early epidemiological studies based on hospital records in Massachusetts suggested that Black and Latino communities are disproportionately affected by COVID-19.[15,16] That is, low-income Black and Latino populations may have more chances to be exposed to the virus due to several factors: (1) they are likely to live in densely populated areas, like a multi-household house; (2) many are not able to work from home; and (3) paid sick days are uncommon.[17,18] In addition, it has been reported that other factors, like population density, median house income, mean household size, the proportion of foreign-born noncitizens, and the share of food service workers, can contribute to the disproportionate burden of COVID-19 infection.[19] Yonker et al.[20] addressed that children can be a potential source of COVID-19 spread despite lack of symptoms, based on medical records of pediatric patients (aged 0-22 years) enrolled at Massachusetts General Hospital. However, these results are analyzed based on scarce data at the time of the initial COVID-19 pandemic and thus require further research.

**Drivers of Heterogeneous Transmission**

This variation in the history of disease transmission takes place against a background of significant variations between different towns in measures of population density and median family income. High rates of transmission are associated with high population density and low median family income, Figure S1. These two factors combined explain 56% of the variability among towns in COVID-19 accumulated cases.

Mechanistically, the high population density limits the potential for social distancing favoring disease transmission. The low median family income is consistent with jobs that may not offer the luxury of working from home, and with the use of modes of public transportation that may enhance exposure to the virus.

The two drivers are related, and both are associated with poor socioeconomic conditions. Low median family income often forces families to live in neighborhoods with high population density where the median home price is likely to be more affordable. As a result, COVID-19 transmission is enhanced under relatively poor socioeconomic conditions.

**COVID19 and Ethnicity**

For historical reasons, the ethnic composition of any town reflects the same two factors, population density and median family income, with a high fraction of Blacks and Hispanic ethnic groups living in towns that are characterized by high population density and low median family income, Figure S2. These two factors combined explain statistically 26% of the observed variability among towns in the combined fraction of these two ethnic groups.

Combining these two trends: disease transmission connection to poverty and historical connections of ethnic composition to the same phenomenon may explain the high rate of transmission of COVID-19 among Blacks and Hispanics ethnic groups, Figure S2.

#### Discussion

The main drivers of the heterogeneity in disease transmission at this scale of towns seem to be differences in median family income and population density. Since the ethnic composition of the same towns reflects these two drivers, the resulting pattern of disease transmission has an ethnic dimension. Hotspots for transmission seem to be disproportionally in urban centers, with relatively high populations of Hispanics and Blacks.

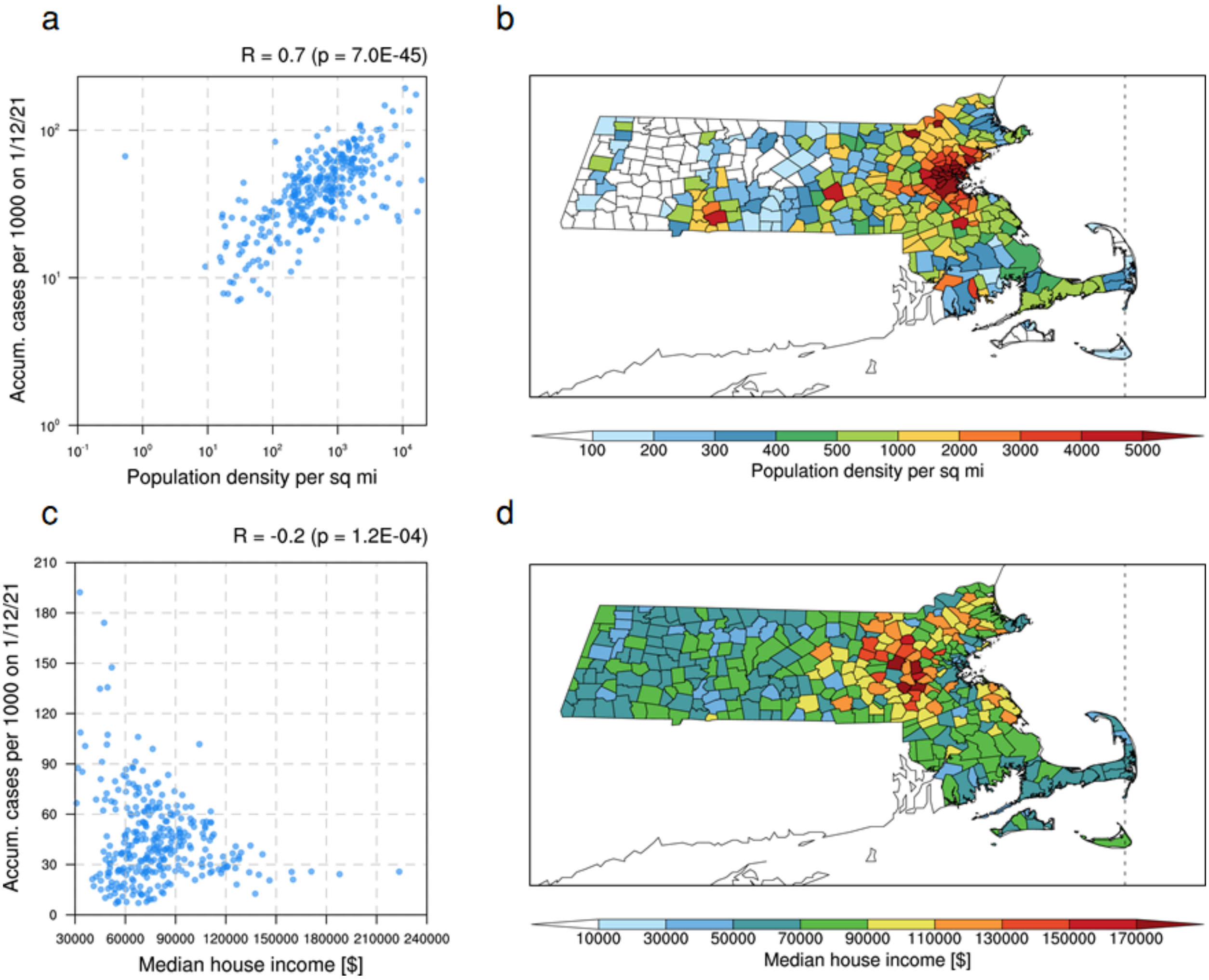


Figure S1. Population density, median house income, and COVID-19 prevalence across towns within the state of Massachusetts. Accumulated confirmed COVID-19 cases per 1,000 people on January 12, 2021, against population density (per square mile) (a) and median house income ($) (c). Spatial distribution of population density (b) and median house income (d). Values indicated on top of (a, c) represent the correlation coefficient (R) and p-value (p).

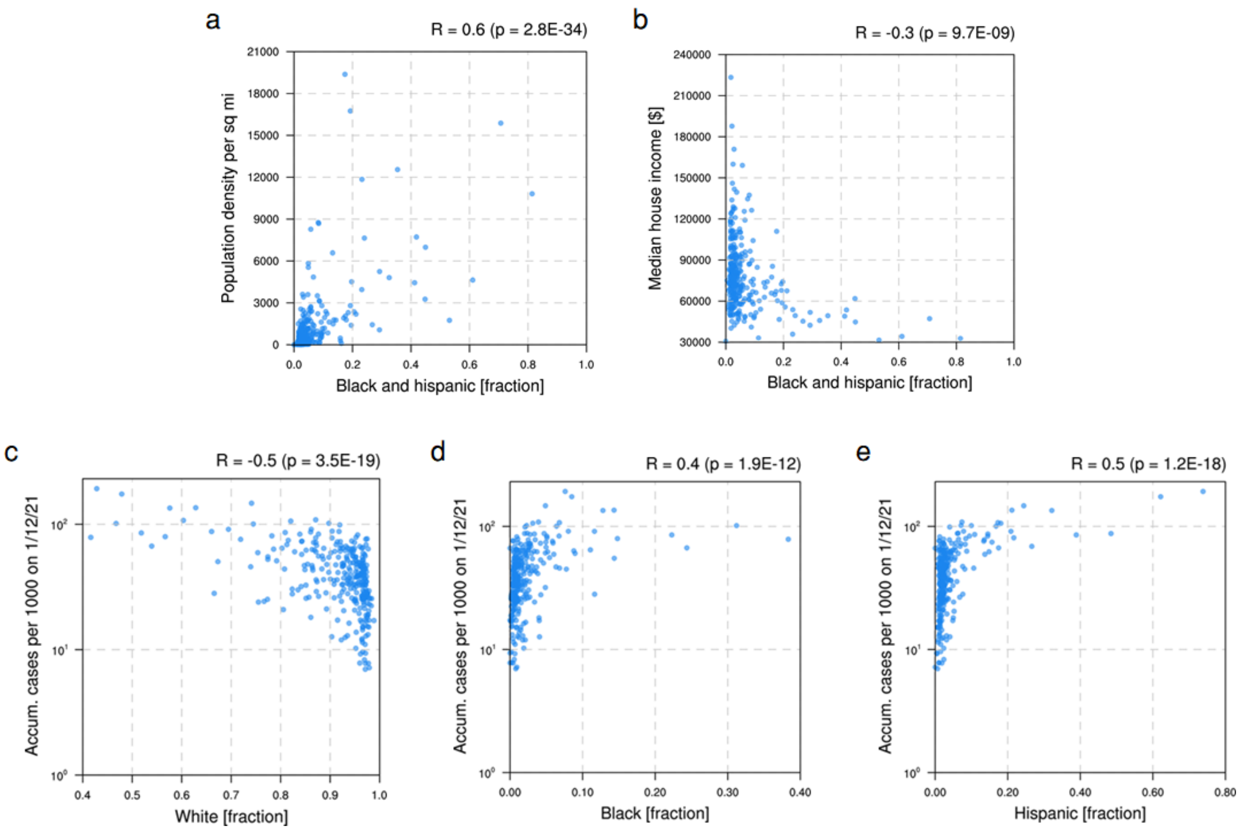


Figure S2. Demographic factors and COVID-19 prevalence across towns within the state of Massachusetts. Fraction of Black and Hispanic ethnicity against population density (per square mile) (a) and median house income ($) (b). Accumulated confirmed COVID-19 cases per 1,000 people on January 12, 2021, against the fraction of White (c), Black (d), and Hispanic (e). Values indicated on top of each plot represent the correlation coefficient (R) and p-value (p).
